# Staff perspectives on conversations about romantic/intimate relationships in mental health social care services: a qualitative interview study

**DOI:** 10.64898/2025.12.02.25341456

**Authors:** Angelica Emery-Rhowbotham, Helen Killaspy, Sharon Eager, C Joanna, Brynmor Lloyd-Evans

**Author notes:** Corresponding author (BLE).

## Abstract

Romantic/intimate relationships are an important part of most people’s lives, including people with mental health problems. However, people with mental health problems continue to have little access to support around romantic/intimate relationships. This study aimed to investigate social care staff perspectives on supporting people with romantic/intimate relationships. We conducted 15 qualitative interviews with mental health social care staff to explore their views on supporting people using services with desired romantic/intimate relationships, facilitators and barriers to these conversations, and strategies to offer support in this area. We purposively recruited staff working in a range of social care roles and with varied demographic characteristics. Interviews were analysed using reflexive thematic analysis. We identified three primary themes. These related to (i) whether romantic/intimate relationship support should be offered; (ii) how romantic/intimate relationship support should be offered; and (iii) whether social care staff are the right people to offer this support. Overall, participants felt that romantic/intimate relationship support is important for the quality of life of people accessing services. However, they noted a lack of resources and training relating to romantic/intimate relationship support, and discussed some safeguarding concerns. Findings highlight the need for clear organisational policy and training to address staff concerns, and research to understand the perspectives of people using services on romantic/intimate relationship support.

**Teaser Text:** Most people value romantic/intimate relationships in their lives, including people who use mental health services. While people with mental health issues often report that they want support with relationships, they often do not have access to it. This study sought to understand why, by interviewing 15 mental health social care staff about their views on providing support around romantic/intimate relationships to people who use social care services. Staff generally agreed that relationship support is important for helping those who use mental health services to feel empowered and improve their quality of life. However, they expressed concerns about a lack of resources and training available to help them offer romantic/intimate relationship support. They also discussed ethical worries, including wanting to protect vulnerable people and what to do when working with people who have an offending history. Accordingly, organisations should develop clear policies and practical training to help staff gain skills and confidence in providing this support. There is also a need for future research to find out more about what service users themselves want and need in this area.

## Introduction

Romantic/intimate relationships can be defined as those involving romantic love, physical intimacy or sexual activity, and are characterised by the World Health Organisation as “a central aspect of being human” (World Health Organisation, 2006). Most people with serious mental health problems value romantic/intimate relationships and see them as a key facilitator and indicator of recovery (Boucher et al., 2016). Satisfaction with one’s relationship status is associated with better mental health and wellbeing (Boysen & Isaacs, 2022). Romantic loneliness, more than family or social loneliness, is a risk factor for suicidal behaviours (McClelland et al., 2023). People with mental health problems commonly report challenges in forming and maintaining romantic/intimate relationships (Forrester-Jones et al., 2024). For instance, for those with psychosis, satisfaction with their sex life was the lowest rated of all life domains (Laxhman et al., 2017). Studies have found that about two thirds of people with serious mental illness are single (Thornicroft et al., 2004; Hughes et al., 2020). Negative, stigmatising attitudes from others can form a barrier to developing fulfilling romantic/intimate relationships for people with mental health difficulties. Surveys of public attitudes indicate that mental illness is seen as a “dealbreaker” for many, leading them to reject a potential romantic partner (Boysen et al., 2019). People with mental health problems may also be viewed as more “sexually exploitable” than others (Boysen & Isaacs, 2022). This may contribute to the high rates of sexual violence experienced by people with mental health problems, especially women (Kaul et al., 2024). Despite their importance for many, people who use services often experience staff as uninterested in their intimate relationships (White et al., 2021). This has been characterised as the “institutional silencing of sexuality” in mental health care (Urry et al., 2022).

Two recent systematic reviews summarise views on needs for support with romantic/intimate relationships in mental health care, from the perspectives of service users (O’Connor et al., 2025) and staff (Wu et al., 2025). O’Connor and colleagues (2025) collated findings from 17 primary qualitative interview studies with people using mental health services. Most participants indicated a desire for discussion and support with romantic/intimate relationships to be offered in mental health services. However, many also reported staff reticence and awkwardness around this topic. They identified poor continuity of care in services as a barrier to developing the secure therapeutic relationships with staff which could make conversations about romantic/intimate relationships feel more comfortable. Notably, most included studies were conducted in psychiatric inpatient settings, so the applicability of findings to community and social care settings is unclear.

Wu and colleagues (2025) included 24 qualitative interview studies in their review of staff’s perspectives of service users’ needs for support with romantic/intimate relationships. Participants reported personal and organisational barriers and reservations around discussing people’s wishes and needs regarding romantic/intimate relationships. These included staff’s lack of confidence in their own capability, other perceived higher priorities for care and support, uncertainty about the limits of their professional role, and a lack of organisational support or training. As with studies of service users’ views (O’Connor et al., 2025), studies with staff were disproportionately conducted in inpatient and forensic settings and none specifically focused on the views of social workers or social care staff.

As identified in a recent survey exploring the views of mental health care staff on offering romantic/intimate relationship support (Author’s Own, 2025), social workers and social care staff are potentially well-placed to support people with mental health difficulties with developing positive romantic/intimate relationships. This clearly aligns with the British Association of Social Workers’ professional code of ethics to “enable all people to develop their full potential, enrich their lives, and safeguard people who may be at risk of harm” (BASW, 2021). Mental health social care is particularly valued for being holistic in its understanding of individuals’ support needs (Dunthorne, 2022). Yet there is an evidence gap regarding the views and experiences of social workers and social care staff on supporting people with their needs and wishes in relation to romantic/intimate relationships. This study aimed to address this gap in knowledge.

## Methods

### Research questions

This study addressed the following research questions:

1. What are the perspectives of mental health social care staff on supporting people using services with achieving desired romantic/intimate relationships?
2. What helps and hinders staff to have conversations about romantic/intimate relationships?
3. What strategies can staff use to support people with their wishes and needs for romantic/intimate relationships?

### Design

This qualitative study involved semi-structured interviews with social workers and social care staff involved in providing care and support to people using mental health services. Data were analysed using reflexive thematic analysis (Braun et al. 2018; Braun & Clarke 2019). This study was approved by the UCL Research Ethics Committee on 22^nd^ June 2023 (ref 20217/002).

### Participants and Setting

We sought to interview staff from a variety of mental health social care (MHSC) roles in England, who provide care and support to adults living with mental health conditions. This included social workers, support workers, peer support workers, and other roles in the social care sector, for example residential care home managers. Mental health staff from other professional disciplines were excluded, as were social workers and social care staff who worked primarily with client groups other than adults with mental health conditions. We also excluded any staff who worked in a specialised relationship counselling, sexuality or gender identity service, as this study wished to focus on the views and experiences of staff providing general mental health social care. We sampled purposively to achieve diversity in participants’ demographic characteristics (age, gender, ethnicity and religion), their work role, service setting, and their level of experience. We aimed to recruit 15 participants in total to hear a range of views and experiences. This also aligns with guidance for adequate qualitative interview sample sizes (Guest et al., 2006; Hennink et al., 2016).

### Materials

We developed an interview topic guide informed by issues raised in two previous studies undertaken by our research group: a focus group study with residential social care staff and residents about meeting residents’ social inclusion needs (Eager et al., 2023), and a recent survey paper which explored mental health staff perspectives in offering intimate relationship support (Author’ s Own, 2025). Interviews began by asking the participant to provide demographic information. Participants were then asked nine open questions, with various prompts attached. These questions included asking about whether wanting a romantic/intimate relationship was a common need among the people using their service, whether they thought romantic/intimate relationship support was appropriate in their job role, what the barriers and facilitators are to having conversations about intimate/romantic relationships, and ways to potentially support people using their services with this need. Please see Supplementary Material for the complete topic guide.

### Procedures

#### Recruitment and consent

Our recruitment procedure included emailing a list of professional contacts known to the research team, collated using their professional networks. This included national networks for social workers, such as Think Ahead and the British Association of Social Workers, national voluntary sector organisations providing mental health and social care, such as Rethink, and a number of local supported housing providers and peer worker networks. These contacts were also asked to share the recruitment information with others in their organisation, in a snowballing sampling approach.

Potential participants contacted the research team by email. The lead researcher then confirmed their eligibility through email or phone discussion. They were sent a participant information sheet and offered the opportunity to ask any questions about the study. Those still interested were sent an invitation for an online meeting through Microsoft Teams. Participants were offered the chance to ask questions about the study, and prior to starting the interview, a consent form was read out by the researcher. Participants were asked to verbally confirm their understanding and agreement to each item and their consent to take part.

#### Data collection and management

Interviews were conducted over a period of 18 weeks (13/10/2023 to 09/02/2024) and lasted up to one hour. Once completed, the Microsoft Teams video recording was converted to an audio file, which was sent to a UK-based professional transcription company. Transcripts were checked for accuracy and any potentially identifying text was anonymised. De-identified transcripts were stored in UCL’s secure online folders. Audio-files of interviews were deleted once analysis was complete.

### Analysis

We used a reflexive thematic analysis approach (Braun et al. 2018, Braun & Clarke 2019). Our approach to generating codes and themes was primarily inductive, driven by the data, although analysis was also informed by our main research questions. Analysis was led by AER. She conducted all interviews, checked the transcripts for accuracy, then re-read them all to achieve data familiarisation. Next, transcripts were uploaded onto NVivo software and were line-by-line coded into ‘meaningful’ units of text. These codes were reviewed with the research team and suggestions for grouping codes into themes were discussed. AER then developed a first set of themes. These were reviewed by the research team in an iterative process with reference to interview transcripts. This involved considering any interview text not adequately captured by existing themes or potential reorganisation or renaming of themes and subthemes, which were revised and defined accordingly. AER then drafted the final report, which was revised in collaboration with the research team.

### Researcher positionality and reflexive statement

Lead researcher AER is a postgraduate researcher from a psychology background. Other authors are researchers from social work (BLE), psychiatry (HK), lived experience (JC), and psychology backgrounds (SE). This paper forms part of a developing programme of research in our group about supporting people with mental health difficulties in developing and sustaining desired intimate relationships. We share a belief that this an important topic which is neglected in research and practice. We acknowledged the risks of this position making us sceptical by default about participants’ concerns about the appropriateness or feasibility of talking to people using services about their needs for intimate relationships. We sought actively to recognise if this appeared to be affecting our approach to analysis and raise with colleagues if apparent in their contributions. We also acknowledged our limited cultural diversity in the author team, and sought to review transcripts carefully so as not to miss text where participants discuss cultural considerations. As well as frequent and iterative discussions in the author team, AER made reflexive notes immediately after each interview and during analysis, to encourage and prompt the team to think about positionality in relation to the data and how this may influence the analytic process.

## Results

### Participant characteristics

We interviewed 15 social care staff in total. All participants but one were female and the majority were white. Most did not adhere to a religion, and most had spent more than five years working in mental health services. Staff roles included social workers, support workers and peer support workers, among others. Over half worked in supported accommodation services. See Table 1 for a full breakdown of participant characteristics.

**Table 1.**
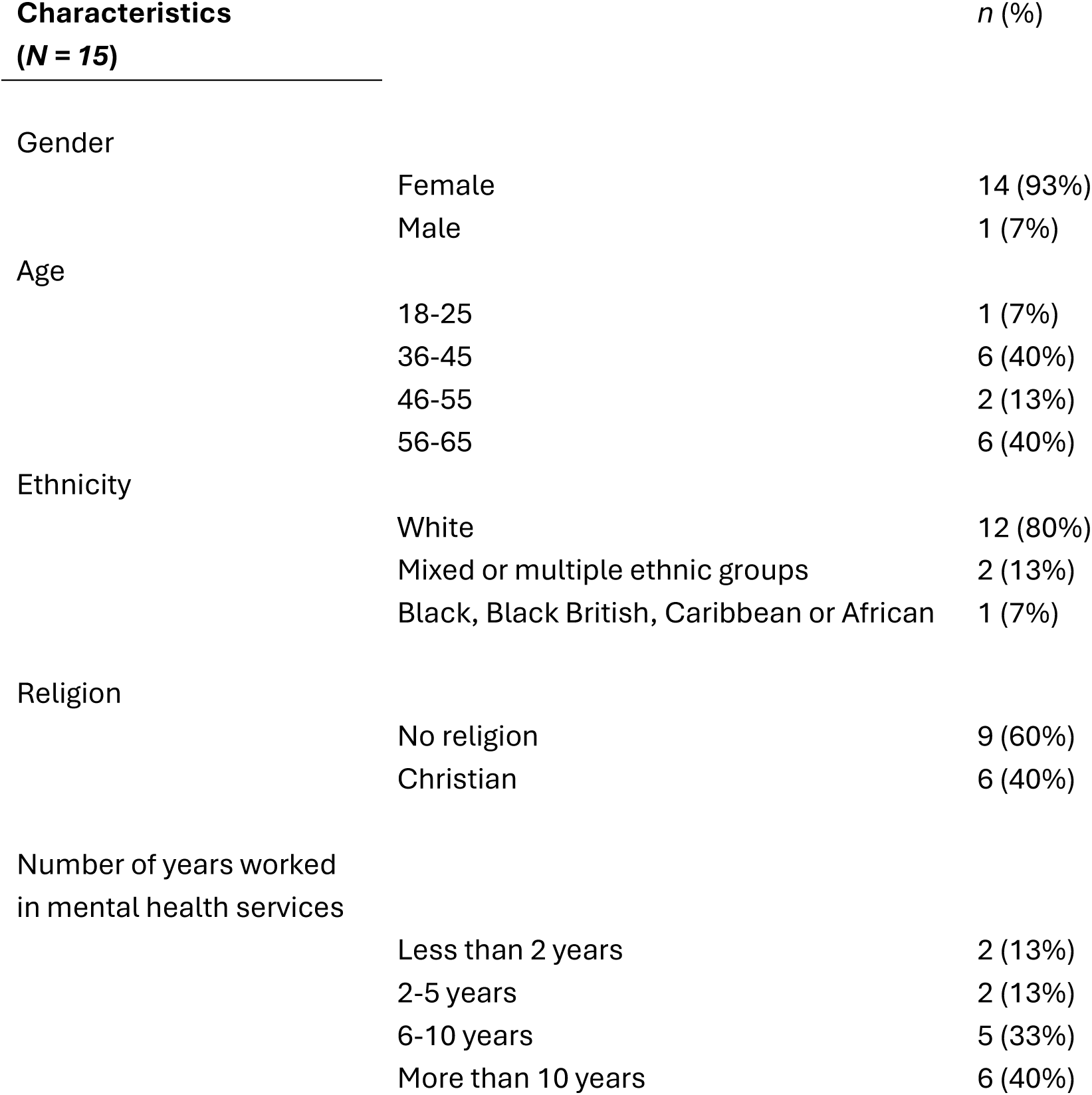
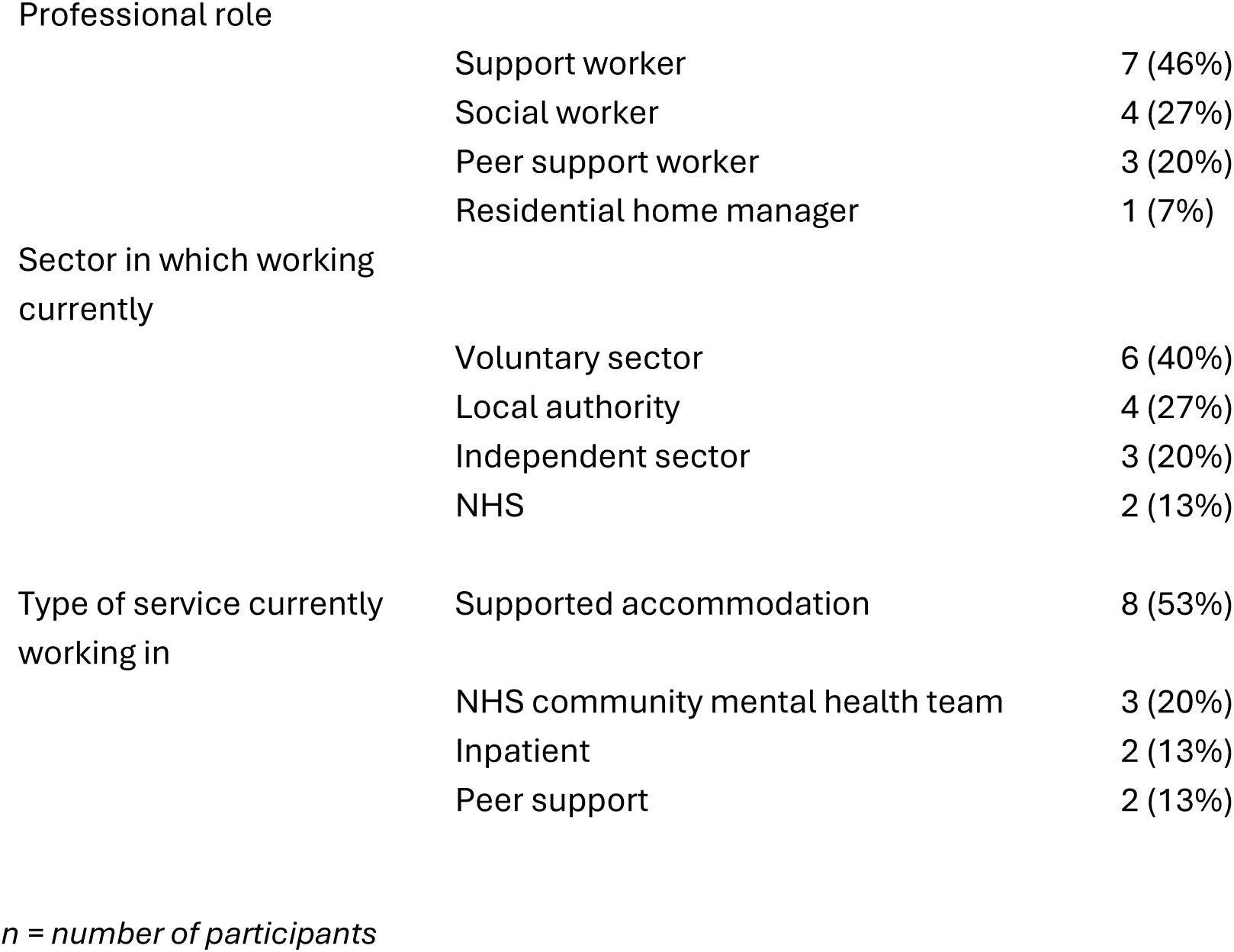
Participant characteristics.

### Overview of themes

We identified three main themes: ‘should we be doing this?’; ‘how should we be doing this?’; and ‘am I the right person to do this?’ (See Table 2 for a summary of themes and sub-themes).

**Table 2.**
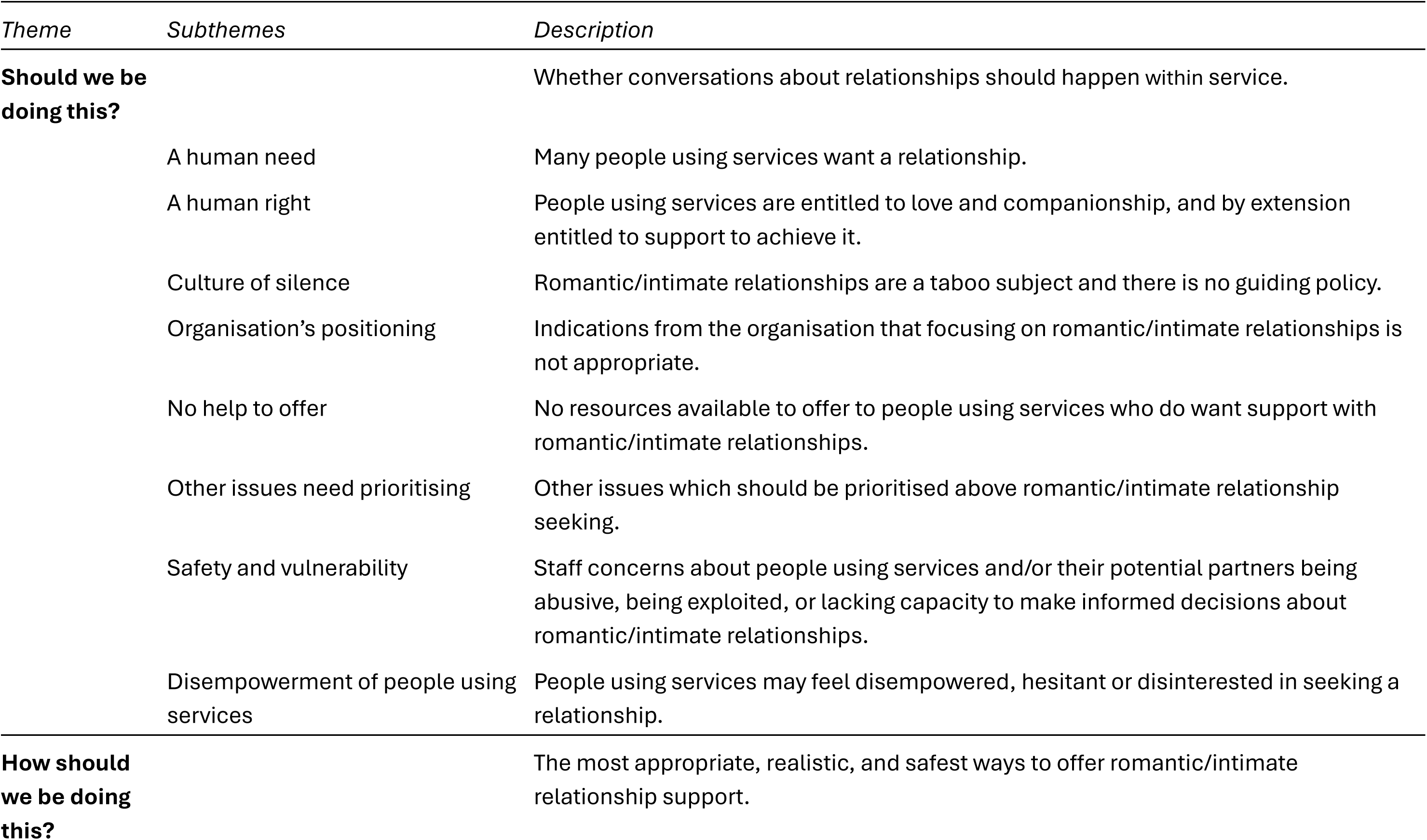
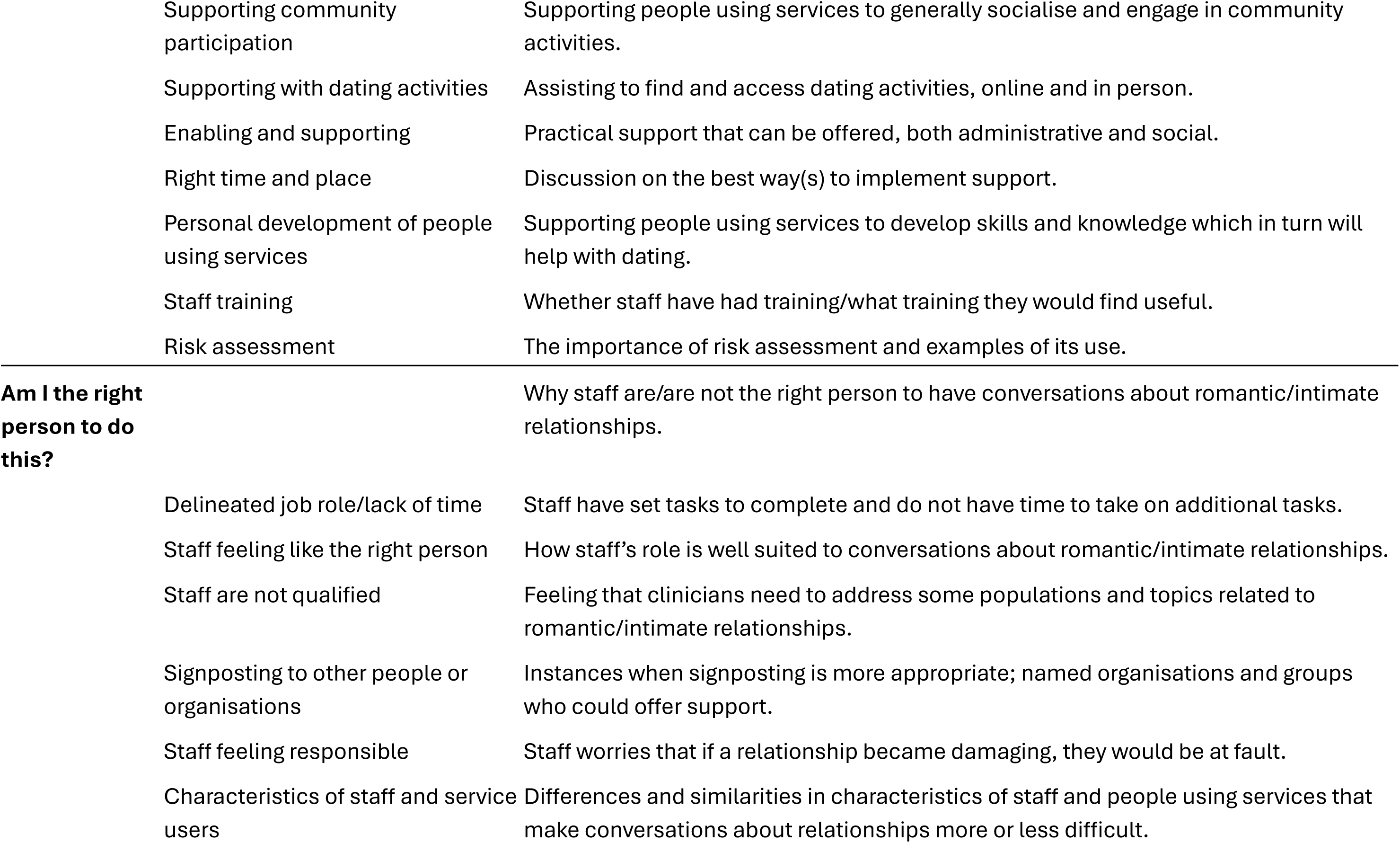
Summary of themes and subthemes.

#### Should we be doing this?

This first theme referred to whether social care staff should be having conversations about romantic/intimate relationships with people using services. Generally, participants felt that intimacy was a basic human need and therefore these conversations should happen, and that such relationships may offer specific benefits. These included providing companionship, combatting loneliness, and an opportunity for people using services to express their sexuality. Also discussed was the idea of relationships being an external signal of “normalcy”:

> *“They already think of themselves as being different because of their mental health. So, having a relationship means that they’re not unnormal or they can be loved … So yes, I would say it’s quite a big thing.” [Participant 4, service manager, supported accommodation].*

Similarly, participants discussed intimacy as being ‘a human right’. They suggested that people using their services should be entitled to love and companionship the same as anyone else, and therefore they should be able to discuss this with staff as an important aspect of life. Some participants described that people using services may have lost a sense of this due to their mental health problem.

> *“Sometimes they come across as forgetting that they also have needs as individuals for intimacy. So I keep on reminding them that everybody is entitled to companionship and love. But they don’t seem to believe that.” [Participant 5, health and social care worker, supported accommodation].*

Some participants also discussed their duty to provide support with relationships as part of the holistic approach encompassed by their social care role:

> *“We haven’t got any specific part of their lives we’re helping with. It’s kind of an all-round support. So social connection and romantic relationships are a big part of people’s lives. So I think definitely it should be part of our role.” [Participant 1, support worker, supported accommodation].*

However, despite most participants’ endorsement for the principle of offering support with romantic/intimate relationships, many also expressed ambivalence about whether this could or should be offered in practice. Staff discussed feeling hesitant about encouraging people to talk on this topic due to a lack of time, resources, and competing priorities in their provision of care. It was also proposed that the people they supported needed to be relatively stable in their mental health and social functioning before any support was offered for romantic/intimate relationships:

> *“I don’t think somebody has to be the finished article to pursue relationships, but I do think… somebody needs to at least have stable housing and be able to meet their own basic needs, otherwise I just think they’re not in an appropriate place to have a conversation like that. It could actually cause quite a lot of stress.” [Participant 9, support worker, supported accommodation].*

Similarly, some staff discussed concerns about romantic/intimate relationships having the potential to destabilise people’s mental health:

> *“There is a risk that you become so involved in the other person that you [the person using services] end up putting their needs first before your own”. [Participant 6, support worker, supported accommodation].*

Others proposed that seeking intimacy through friendship should take priority above romantic or sexual intimacy, due to the potential of romance arising out of a friendship or platonic connections being seen as sufficient:

> *“At the end of the day, what you need is good friends, good connections around yourself, that’s the most important thing.” [Participant 3, support worker, supported accommodation].*

Some participants felt that the subject of supporting people with finding romantic/intimate relationships was taboo within mental health services, which meant it was rarely discussed with colleagues or brought up by managers. This was echoed by a lack of organisational policy on the topic to guide staff:

> *“It doesn’t get mentioned, we talk about everything … about how you ask someone whether they need help with toileting … potentially embarrassing questions, but we don’t really talk much at all [about romantic/intimate relationships] amongst social workers. It’s not really something that is discussed or brought up.” [Participant 8, social worker, community mental health team].*

In addition, participants described that some of the people using their services appeared disempowered about finding a relationship because of past negative experiences, stigma, or due to having a history of relationship-related offences:

> *“When you get to the more intimate things like, “Do you have any regrets about relationships?” they are much more sensitive questions that could potentially upset a lot of people … Like these two women … had been extremely violent to ex-partners and had really delusional beliefs about those men. And that would have been … unhelpful and potentially dangerous to ask because of … the pain and anger.” [Participant 13, social worker, inpatient service].*

Some staff felt that users of their services may not be interested in the topic because of the impact of their medication on their energy or libido:

> *“It’s the medication. Patients feel lethargic. They don’t give a damn. If they don’t give a damn, why would they bother to have a relationship?” [Participant 3, support worker, supported accommodation].*

Others emphasised that some people using services were content without a relationship:

> *“Some people are … happy as they are and don’t really miss it, and then it’s not for me to tell you actually, you should have someone.” [Participant 8, social worker, community mental health team].*

Another factor was safety and vulnerability. Some participants discussed safeguarding concerns, such as people’s potential vulnerability to exploitation from others and the possibility of them becoming involved in an abusive relationship:

> *“Equally, you know that they’re really vulnerable and could be really easily coerced so it can be really difficult with safeguarding elements in mental health around this stuff.” [Participant 11, peer support worker, community crisis team].*

Conversely, some staff highlighted situations where a person using services might cause harm to a potential partner, for instance through exploitation:

> *“For example [a person using services] might need money for drugs. So they use intimate relationships to get that money. So they either do street working or they try to take advantage of the individuals … to get what they want.” [Participant 5, health and social care worker, supported accommodation].*

Relatedly, participants discussed individuals’ capacity to make potentially unwise, yet informed decisions about relationships, and limitations in staff’s ability to mitigate risk:

> *“It’s going back to capacity. They’re allowed to make those unwise decisions. Even if I can’t stand the person that they’ve been dating, I’ve still got to support them to find the routes and the answers that they want themselves.” [Participant 2, nursing home manager, supported accommodation].*

#### How should we be doing this?

In this theme, participants described the most helpful, achievable, and safest ways for staff to offer people using services support with romantic/intimate relationships. A common suggestion was supporting people to access social groups and community activities to widen their social circle and develop friendships, which could then naturally develop into a romantic/intimate relationship:

> *“What I’m doing is supporting people to socialise more … [when] they’ve got the opportunity of having friendships with people of the gender they’re interested in … things develop a bit more naturally.” [Participant 8, social worker, community mental health team].*

Participants also discussed the potential of direct support, for instance by enabling access to dating activities and dating apps. Staff generally agreed that these would be appropriate methods of support, provided it was instigated and led by the person using services:

> *“Yes, I think that is appropriate, as long as it’s led by the person and what they need. It wouldn’t be appropriate to start telling somebody that they should use dating sites. But I think it’s appropriate to give somebody the options of how they might go about meeting someone and offer to support them with that.” [Participant 9, support worker, supported accommodation].*

Potential issues, such as coming across staff members on dating apps, were discussed, as was the importance of conducting a risk assessment prior to dating app use. Specific activities which could enable and support people using services to find a relationship were also described, such as organising travel arrangements to dates, accompanying/chaperoning them to dates, and providing emotional support and reassurance to prepare them for social engagements.

Although some staff raised the possibility of people using services hiring a sex worker to attend to their sexual needs, in general this was not felt to be an appropriate method of support. This was due to its illegality and concern for the safety of the sex worker. As one participant described:

> *“I think it didn’t help that [this particular] client … is someone with a forensic history who poses a risk to others. I think the case law [said] that we do have a duty to the general public and we cannot encourage him to do something which would then put a potentially vulnerable sex worker at risk.” [Participant 8, social worker, community mental health team].*

Several staff members felt that they had a role in supporting people’s personal development through encouraging skills such as cooking, cleaning, and personal hygiene. They felt this could facilitate people using services’ feelings of capability for attempting social challenges and their preparedness for engaging in a romantic/intimate relationship. Participants also recognised that having low self-esteem was a potential barrier for instigating a romantic/intimate relationship, and that building self-confidence may help those using services to pursue this. Furthermore, participants reported that providing healthy relationship education, such as how to create and maintain safe boundaries, was an important part of their role in supporting people who were seeking a romantic/intimate relationship.

Regarding the best ways to implement support, staff often described the need for an individualised approach when supporting people seeking a romantic/intimate relationship:

> *“Just because they’re all young or whatever, doesn’t mean that they’re all going to want to go out and get married next week. So it’s important to treat them all as individuals … because they’re all going to have a different idea, or whether they want it at all.” [Participant 6, support worker, supported accommodation]*.

Staff also spoke of the importance of using clear communication and building trust before discussing intimate relationships with people using services:

> *“If the person is more reserved and it feels more intrusive for me to ask questions altogether, I’m not going to go there.” [Participant 8, social worker, community mental health team].*

Participants also highlighted the importance of boundaries in the content of these conversations:

> *“I guess if somebody wants to have conversations which are too much about sex and their sexual lives rather than relationships, then we wouldn’t think that’s very appropriate for us to discuss that with them.” [Participant 10, peer support worker, community mental health team].*

Staff also reflected on instigating conversations about relationships. Most felt that it should be the client’s responsibility to signal their desire for support. They also noted that when people using their service did bring up the topic of relationships, these conversations often arose out of discussions about loneliness:

> *“A lot of the time when we start to work with somebody new, they just mention feelings of isolation, whether that’s friendships, relationships, family…” [Participant 15, peer support worker, community mental health team].*

Otherwise, some staff highlighted that intimate relationships might be discussed after a person using their service had acted or spoken in an inappropriate way:

> *“It’s usually that someone has said something that is really crass and cringeworthy or someone’s touched somebody’s bottom. It comes out.” [Participant 7, support worker, supported accommodation].*

Participants also described topics they would like to receive training on. This most often related to wanting clear guidance on what type of support is appropriate for them to provide and the ways in which it should be offered:

> *“I would like guidance around what we can do practically, so whether we can be signposting people to dating nights, whether we can be helping people set up online profiles… I have never known whether that is an okay thing for us to do.” [Participant 1, support worker, supported accommodation].*

Several participants expressed a need for specific training to help them to feel capable and confident about addressing the topic of romantic/intimate relationships:

> *“I don’t think it’s taboo or inappropriate or whatever. Even if they were talking about sex, it’s not necessarily that it’s not appropriate, it’s just that I don’t have the training to deal with it so I would have to talk to my supervisor or signpost.” [Participant 10, peer support worker, community mental health team].*

Another suggested topic for staff training was how diagnoses and medication can affect romantic desire and libido. Some staff were keen to learn more about these issues for their own understanding and so that they could help to educate people using services on these topics:

> *“So when [a person using services] can have a proper relationship, [they] understand more the impacts that can hinder relationships, from lethargy, lack of libido, all those things because if … the other person doesn’t feel desired, then they reject them.” [Participant 3, support worker, supported accommodation].*

Staff also noted that training needed to be inclusive of diverse sexual and gender identities:

> *“I’d also like some input on dealing with issues around sexuality and gender issues. I think that would be really important.” [Participant 7, support worker, supported accommodation].*

#### Am I the right person to do this?

In this theme participants discussed personal factors, such as how their own demographic characteristics, life experiences, and personal views influenced their confidence and attitude towards having conversations about romantic/intimate relationships with people using their service.

> *“I think staff members, yes, would feel a little bit uncomfortable having these conversations, especially if it maybe led on to a conversation about sexuality. I think staff maybe think, ‘Well if I start this conversation, where could it end? Why ask if they want a girlfriend and suddenly we’re talking about masturbation?’ I would feel out of my depth.” [Participant 9, support worker, supported accommodation].*

Participants also discussed some personal attitudes, noticed in themselves or in other staff, which may affect whether they offer romantic/intimate relationship support:

> *“I think it’s difficult for support workers to maybe see our residents as people with a sexuality and needs in that area. Whereas they’re maybe more seen in the context of their mental and physical health issues. I think there’s a barrier there to seeing somebody with a disability. I think the automatic response of many people is to not see that that person is a sexual being.” [Participant 9, support worker, supported accommodation].*

Some participants reflected on how having their own experience of mental health issues influenced their approach. One reflected that this experience made them worry more, as they knew how easily recovery could be impacted negatively by intimate relationship difficulties. Another reflected that it made them more likely to want to provide support to people in this area:

> *“I have mental health problems, so I have seen the stigma around mental health first hand. I would hate somebody to say I’m not entitled to companionship or love or intimacy because I have some difficulties. I would hate that to be the case for my residents as well”. [Participant 5, health and social care worker, supported accommodation].*

Similarly, the personal boundaries of staff members were noted. Some felt that they would always be uncomfortable talking about romance and sexuality due to their own personal experience or upbringing. Others reflected on the potential to feel responsible if they had supported someone to seek a relationship and the experience became damaging:

> *“As much as I would like to feel comfortable doing it, I think that would make me quite nervous … if they were to go out on a date and try and have a romantic connection and it doesn’t go well and then that sets them back in their recovery, yes, I would probably feel quite upset.” [Participant 1, support worker, supported accommodation].*

Staff also noted how demographic differences between themselves and people using services influenced whether they felt comfortable having discussions about finding a romantic/intimate relationship. Participants described feeling wary of causing offence regarding these kinds of discussions:

> *“I would worry more about speaking to someone about romantic relationships or intimacy if they’re from a very different religious background to me because I’m not sure, I wouldn’t want to offend.” [Participant 1, Support Worker, Supported Accommodation].*

Some staff highlighted a particular challenge in supporting people using their service who had a forensic history in regard to romantic/intimate relationships. They felt that this created a particular barrier to discussing romantic/intimate relationships and may bring up feelings of shame for the person using the service if their forensic history was related to partner violence or sexual offending. In these situations, staff also had concern for the safety of potential partners and some felt out of their depth to be able to discuss this topic, due to the complexity of these kinds of situations:

> *“…He has a forensic history. From his personality type, there are concerns, now that he’s in a relationship, of him committing domestic violence against his partner. It needs to be risk assessed, whether it’s appropriate, and whether you are actually able to then support the person to engage in a relationship safely. We do have a duty of care to other people and the general public.” [Participant 8, social worker, community mental health team].*

A few participants reported feeling that they were the right person to discuss romantic/intimate relationships due to their particular role in the team:

> *“I think general discussion around finding a relationship can be done by a support worker. I think we have the ability to discuss [that] with them … and they should feel comfortable talking about that with us because it’s a big part of life.” [Participant 1, support worker, supported accommodation].*

## Discussion

The current study offers new insights into how mental health social care staff can support those using services with romantic/intimate relationships. Overall, three overarching themes describing staff sentiments on this topic were identified: “should we be doing this?”, “how should we be doing this?” and “am I the right person to be doing this?” A sense of ambivalence from staff is evident throughout these themes.

While most participants expressed their belief that this is an important, often neglected topic, they also had several practical and ethical reservations. Staff balanced being empowering versus being protective: for instance, believing that offering support with relationships is important and encompasses holistic psychological support, but worrying that someone’s mental health may suffer when beginning a romantic/intimate relationship. Practical concerns included not having enough time, resources, training, or organisational backing to offer this type of support. Staff flagged additional concerns when discussing this topic with people who had a forensic history, especially those who had committed sexual offences. They also reflected on their hesitation to offer relationship support to those whose demographic characteristics were different to their own, for instance service users of a different cultural background, sexual orientation, or those of a different generation.

Regarding what support could be offered to people with intimate relationships, participants mostly discussed what could theoretically be done rather than support strategies already employed. These included indirect approaches, such as supporting a service user to generally socialise or building their confidence, and direct methods, such as chaperoning service users to dates or assisting them with creating a dating profile. The ethics and associated risks of each of these methods were key considerations.

Most participants expressed a wish for clear organisational guidance about the support they should provide to people who use services with romantic/intimate relationships. Participants also described wanting practical training on how to handle potentially delicate conversations, including education about the impact of psychiatric medication on libido and sexual functioning, and training to understand how to address sexuality and gender diversity.

### Strengths and Limitations

A strength of this study is that the sample represented a broad range of mental health social care roles including social workers and social care staff from Local Authority, health, and voluntary sectors, and those working in varied service settings. The research team included lived experience and practice perspectives, and we strove to work reflexively throughout the interview and analysis process.

A major limitation is the limited gender, ethnic, and religious diversity within the sample. Firstly, with only one male participant out of fifteen, unique insights such as discomfort in supporting young women with intimate relationships were unable to be explored as a broader male experience. Moreover, with 80% of participants identifying as white and all reporting either Christian beliefs or no religious beliefs, the current findings do not reflect more diverse ethnic and religious viewpoints.

Although we utilised purposive sampling to recruit a variety of participant roles and settings, it is likely that staff who were more comfortable talking about this topic were more likely to participate. Consequently, the current findings may overrepresent staff who are comfortable with having conversations about romantic/intimate relationships with service users. Therefore, staff confidence and interest in this topic may be overestimated.

### Implications for research

Our study helps address a gap in knowledge about the perspectives of social workers and social care staff in supporting people with mental health difficulties with romantic/intimate relationships. Further qualitative interview studies are warranted to explore this topic further, in England and internationally. Such studies should prioritise hearing from a diverse group of staff, including men and people from minoritised ethnic and faith communities, who were under-represented in the current study.

Most participants in this study felt that talking to people about their needs and wishes for intimate relationships should be offered by mental health social care. However, staff highlighted training needs and organisational clarity and support to do this safely and confidently. This implies two areas where further research is needed. First, there is a need to develop and evaluate organisational guidance and training for staff in how to initiate and conduct conversations with people using services on this topic. The Supported Loving Network suite of resources developed for the intellectual disabilities sector in social care (Choice Support, 2019), provide a model of how resources and a community of practice can be established. Our study highlights that all staff training should address working with cultural and religious diversity, as well as diversity regarding sexual orientation – areas where our participants expressed a lack of confidence. Our study also suggests that additional guidance may be needed for providing support in this area to people with forensic histories, especially sexual offences, where our participants flagged additional complexities and concerns.

Second, there is a need to develop and test specific programmes of care and support to help people who wish to establish positive intimate relationships. A recent systematic review of interventions to promote intimate relationships in mental illness (Caiada et al., 2024) identified a lack of evidence for programmes aimed at people not already in established relationships (as opposed to couples therapy or similar for those who are).

Two recent French studies have piloted psychological interventions for men with psychosis to talk about romantic relationships (Hache-Labelle et al., 2020, Cloutier et al., 2023), but as yet these lack evidence of effectiveness and may not be deliverable in mental health social care settings. The Tapestry Social Relationships Agency was developed and run in Reading, England in the early 2000s (Proudlock & Hallé, 2006). This comprised educational groups about preparing for relationships, individual coaching, and social group meetings in the community. Tapestry offers an exciting example of how support with intimate relationships can be provided in mental health social care, but it has yet to be evaluated.

### Implications for practice

Most participants in our study endorsed the importance of supporting people with intimate relationships as an integral part of providing holistic care and supporting someone to live a fulfilling life. This is consistent with the concept of promoting personal recovery in mental health care, relating to established recovery dimensions of connectedness, hope, identity, meaning and empowerment (Leamy et al., 2011).

In England, government guidance directs social workers and social care staff to talk to people and assess needs with romantic relationships. Developing and maintaining personal relationships is an eligible need for support under the Care Act (Gov.UK, 2014). Furthermore, the Care Quality Commission guidance from 2020 “Promoting sexual safety through empowerment” (Care Quality Commission, 2020) tasks social care staff with creating a culture where “people and staff feel empowered to talk about sexuality”, as an essential prerequisite to assessing risks and providing any individualised support around sexual safety. There are also statutory limits to what support can be provided by social workers and social care staff. In England, for instance, section 39 of the Sexual Offences Act 2003 prohibits care staff from facilitating contact with a sex worker for vulnerable people (Gov.UK, 2003).

Notwithstanding the current lack of practice resources and evidence-based models of support specific to mental health social care around intimate relationships, we suggest there is a clear practice need and policy imperative for organisations to provide guidance and support to staff in the extent and limits of what is encouraged and acceptable in supporting people using services with romantic/intimate relationships. Our study suggests this is not the norm in current practice. As a minimum, we suggest organisations and service managers should develop a written policy around supporting people with romantic/intimate relationships, and proactively encourage discussion of this area of practice with staff in supervision. This could help break the current “stand off of silence” which is common in practice (Author’s Own, 2025), whereby both staff and people using services feel inhibited from initiating conversations about relationships because they are both unsure what is acceptable to talk about in a service context.

People value mental health social care because it “thinks about my whole life, not just my illness” and social workers and social care staff are sometimes more trusted than mental health NHS services, especially by some marginalised communities (Dunthorne, 2022). Mental health social care services and practitioners are therefore potentially well placed to support people using services with addressing their needs and wishes for intimate relationships. Our study highlights some of the challenges and suggests a gap in current research and practice to enable staff to do this confidently and effectively.

## Supporting information

Supplementary Material 1_Topic Guide

## Data Availability

Anonymised data produced in the present study are available upon reasonable request to the authors

