## Supplementary Material 1_Topic Guide for "Staff perspectives on conversations about romantic/intimate relationships in mental health social care services: a qualitative interview study"

**“Finding a relationship” conversations between staff and service users in mental health social care services: a qualitative staff interview study**

**Qualitative Interview Topic Guide V1, 11/04/23**

Thank you for taking part in this study. This interview will involve questions on the topic of relationships, specifically “finding a relationship” conversations between mental health social care staff and service users. “Finding a relationship” conversations entail conversations where the service user and care provider discuss the prospect of the service user finding an intimate relationship. A relationship may constitute a singular partner, or other type of dynamic such as a non-monogamous or polyamorous relationship. We are interested in conversations staff have with service users who currently have no romantic/intimate relationship, or with service users who would like a new relationship, different from the one they currently have. We would like to hear about the context and content of such conversations in mental health social care settings, as well as any barriers perceived by care providers in having such conversations and what types of support might help. Please do not discuss personal information or details that are specific to any service user, in a way that they might be identified based on what you’ve said.

**1. Is wanting to find a relationship a common need among the service users you support in your work role?**

**Prompts:**

- **What proportion of your service users would you say would like to find a relationship?**

**2. Do you think talking to service users about finding a relationship and trying to help them with this is appropriate in your work role? Please tell us why, or why not.**

**3. If and when you do talk to service users about finding a relationship, how do these conversations come about?**

*Prompts:*

*- who initiates these conversations?*

*- Do they come up in one-to-one settings or in group settings?*

*- Informally, or as part of structured care planning or assessment?*

*- How often are these conversations brought up?*

**4. What barriers, if any, are there to talking to service users about finding a relationship in the service where you work?**

*Prompts:*

*- Job description and work role?*

*- Feeling that you may break professional boundaries or make the service user uncomfortable?*

*- Lack of time/other work demands?*

*- Lack of training or confidence in how to go about this? If so, what does this lack of confidence relate to?*

*- Feeling of responsibility towards service user or concern if something goes wrong?*

*- General lack of support from your organisation?*

**5. Are there some service users with whom you feel it is more appropriate and possible to have conversations about finding a relationship than others? Please tell us why?**

*Prompts:*

*- How well you know the service user already, or strength of rapport?*

*- Service users' age, gender, religion, culture/ethnicity, sexuality?*

*- Service users' capabilities and stage of recovery (e.g. mental health stability, extent of social skills and functioning)*

*- Service users' previous history of trauma, abuse or sexual violence, which may make conversations about relationships more sensitive*

*- Service users' potential vulnerability to sexual or other forms of exploitation*

*- Service users' capacity to give informed consent to a sexual relationship*

**6. What, if anything, can you do to help a service user in your service who wants to find a relationship?**

*Prompts:*

*- Planning where the person can go to meet new people?*

*- Help with confidence building, independence, and empowering them to be more autonomous?*

*- Social skills training?*

*- Educating service users on what constitutes a healthy relationship?*

*- Discussing dating sites and making dating profiles?*

*- Talking to service users about their thoughts and feelings on the subject, and being a listening ear?*

**7. Please tell us about any guidance or training you have had about talking to service users about finding a relationship and assessing and addressing these kinds of personal needs**

*Prompt:*

*- What training, if any, would you like?*

*- What policies, if any, does your organisation have to guide you in this area?*

**8. What other agencies, if any, should be involved with supporting mental health service users with finding a relationship, and how could they help?**

*Prompt:*

*- Please tell us about any organisations which provide examples of innovative practice*

*- Is there a role for collaboration across different organisations and staff groups? If so, who and how would this work?*

**9. Is there anything else you would like to say which would help us understand how mental health social care staff do or could support people with finding a relationship?**

**Demographic information**

1. How old are you?
   - 18 - 25
   - 26 – 35
   - 36 – 45
   - 46 – 55
   - 56 – 65
   - Over 65
   - Prefer not to say

2. Which of the following best describes your gender?
   - Male
   - Female
   - Describe gender with another term (please tell us your preferred term)
   - Prefer not to say

3. Which of the following best describes your ethnic group?
   - White
     - English/Welsh/Scottish/Northern Irish/British
     - Irish
     - Gypsy or Irish Traveller
     - Any other White background, please describe
   - Mixed/Multiple ethnic groups
     - White and Black Caribbean
     - White and Black African
     - White and Asian

    - Any other Mixed/Multiple ethnic background, please describe
- Asian/Asian British
    - Indian
    - Pakistani
    - Bangladeshi
    - Chinese
    - Any other Asian background, please describe
- Black/African/Caribbean/Black British
    - African
    - Caribbean
    - Any other Black/African/Caribbean background, please describe
- Other ethnic group
    - Arab
    - Any other ethnic group, please describe
- Prefer not to say

4. Do you follow a specific religion?
   - Buddhism
   - Christianity
   - Hinduism
   - Judaism
   - Islam
   - Sikhism
   - Other (please specify)
   - No religion
   - Prefer not to say

5. How long have you worked in mental health services?
   - Less than 2 years
   - 2-5 years
   - 6-10 years
   - More than 10 years

6. What is your current job title?

7. Which professional group, if any, do you belong to?
   - Social worker
   - Trainee social worker
   - Support worker
   - Peer support worker
   - Other (please specify)

8. Which of the following sectors do you work in?
   - Local authority
   - Voluntary sector organisation

- NHS
- Private sector organisation

9. What sort of service do you mainly work in?
    - NHS community based mental health team
    - Day service (e.g. day centre, drop-in service, recovery college)
    - Supported accommodation service (e.g. residential service, supported housing, floating outreach)
    - Inpatient service
    - Other (please specify)
    - Prefer not to say
